# Total-body [^11^C]carfentanil PET: liver-brain axis in methadone vs. buprenorphine treatment

**DOI:** 10.1101/2025.10.06.25337456

**Authors:** Elizabeth J. Li, Corinde E. Wiers, Chia-Ju Hsieh, Hsiaoju Lee, Alexander Schmitz, Anthony J. Young, Timothy Pond, Joseph Chang, Erin K. Schubert, Robert H. Mach, Henry R. Kranzler, Jacob G. Dubroff

**Author notes:** Equal contribution (Elizabeth J. Li, Corinde E. Wiers). Corresponding Authors: Jacob Dubroff, M.D., Ph.D., Department of Radiology, Perelman School of Medicine, 3400 Spruce Street, 1 Silverstein, Philadelphia, PA 19104, Henry R. Kranzler, M.D., Department of Psychiatry, Perelman School of Medicine, University of Pennsylvania, 3535 Market Street, Suite 500, Philadelphia, PA 19104, United States.

## Abstract

**Purpose:** Examine the relationship between brain mu opioid receptor (MOR) availability and liver activity in opioid use disorder (OUD) patients treated with methadone (MET) or buprenorphine (BUP).

**Methods:** 10 OUD patients, 5 treated with MET and 5 with BUP, as well as 13 healthy controls (HCs) underwent total-body PET [^11^C]carfentanil (CFN) imaging. MOR availability was quantified via distribution volume ratio in key MOR-rich brain regions, and hepatic distribution volume (Vt) was used as an index of systemic tracer pharmacokinetics. Serum drug levels were measured in the OUD patients.

**Results:** MOR brain availability was significantly lower in the OUD groups relative to HCs (p<0.001), more so in BUP-treated (mean MOR difference=39.9±15.9%, Cohen’s *d*=3.35) than MET-treated (mean MOR difference=14.2±9.9%, *d*=1.90) patients. Central MOR availability was inversely related to serum drug levels, linearly in MET treated (R²=0.83) and logarithmically in BUP treated (R²=0.96), consistent with distinct receptor occupancy dynamics. Hepatic Vt differed across groups (p<0.003) with levels in both OUD groups lower than in HCs (p <0.05). Hepatic Vt was strongly associated with brain MOR availability (R^2^=0.68) across serum drug levels in MET patients (R^2^=0.63). This relationship was decoupled of this relationship in BUP patients (R^2^=0.30), and absent in HCs.

**Conclusions:** Drug-specific, brain-liver pharmacokinetic profiles were observed in OUD patients treated with MET versus BUP. While MET patients demonstrated coupling between hepatic tracer kinetics and brain receptor occupancy, BUP patients did not. These findings demonstrate that drug-specific brain-body pharmacokinetics may underscore observed clinical differences in opioid agonist effects in OUD.

## Introduction

Opioid use disorder (OUD) represents a global epidemic [1–3]. Methadone (MET), a full mu-opioid receptor (MOR) agonist, and buprenorphine (BUP), a high-affinity partial MOR agonist, are both effective first-line pharmacotherapies for OUD [4]. Despite their clinical efficacy, the drugs show important differences in treatment retention, dosing, safety profiles, and accessibility, with MET typically administered under closely supervised conditions and BUP more commonly prescribed in office-based settings [5–8]. Although both medications reduce illicit opioid use, their distinct pharmacological properties suggest that they differentially engage MORs across brain circuits, yet the neurobiological basis for these differences remains incompletely understood.

We sought to elucidate the pharmacological differences between MET and BUP using positron emission tomography (PET) brain imaging of opioid receptors, which has yielded key insights into MOR behavior [9–14]. Studies using the MOR-selective agonist C-11-labeled carfentanil ([^11^C]CFN), a potent MOR agonist and close analog of fentanyl, have demonstrated dose-dependent relationships between BUP and MOR occupancy, and associations between receptor occupancy and suppression of opioid withdrawal signs and symptoms [11, 12, 14]. Prior PET studies have also shown that opioid agonist treatments modulate receptor availability in humans. Kling et al. [15] reported 19-32% lower opioid receptor availability in multiple MOR-rich brain regions in MET-maintained patients with OUD using [^18^F]cylofoxy PET (a mu- and kappa-opioid receptor agonist), while Melichar et al. [16] observed no differences using [^11^C]diprenorphine, a non-selective opioid antagonist. Meta-analytic and comparative clinical studies indicate that MET and BUP differ in clinical outcomes, with BUP showing lower retention at fixed low doses, while medium and high doses of either medication show comparable efficacy in reducing illicit opioid use. Although MET is associated with higher retention in treatment, it also has a higher overdose risk than BUP [5–8]. Despite these important clinical and imaging findings, direct comparisons of MET and BUP using a selective MOR agonist tracer, and simultaneous assessment of systemic pharmacokinetics, remain unexplored.

The MOR is widely distributed across mesolimbic and striatal circuits [17], where it plays a central role in reward processing [18], affect, stress responsivity, and habit formation [19].

Variation in MOR engagement across these networks may therefore contribute to the differences in clinical outcomes observed between the opioid agonist therapies. However, receptor-level pharmacodynamics represent only one component of drug action. Systemic pharmacokinetics, including absorption, distribution, metabolism, and clearance, also critically shape central drug exposure. MET is primarily metabolized via hepatic cytochrome P450 enzymes, particularly CYP2B6, whereas BUP is primarily metabolized via CYP3A4 pathways [20, 21], suggesting that hepatic processes may differentially influence systemic availability of these medications. Like fentanyl, carfentanil is largely metabolized by the CYP3A4 pathway[22, 23].

Because the relationship between peripheral drug disposition and central MOR availability has not been directly characterized *in vivo*, it is unclear whether systemic pharmacokinetics and brain receptor occupancy operate as independent processes or as a coupled physiological system that differs across opioid agonist treatments. Addressing this gap requires simultaneous quantification of central receptor binding and whole-body pharmacokinetics.

Building on our previous reports[13, 24], we extend the application of total-body [^11^C]CFN-PET to compare the central and peripheral effects of MET and BUP in individuals with OUD. Here, we use the prototype long axial field of field-of-view PennPET Explorer instrument to quantify both brain MOR availability and peripheral tracer kinetics, including hepatic distribution volume (Vt), in individuals with OUD who are being treated with MET or BUP. This approach enables, for the first time, an integrated assessment of systemic and central compartments within a unified imaging framework. We also compare these patients to healthy controls previously scanned under identical conditions to characterize group differences in receptor availability and tracer kinetics.

We hypothesized that (1) both MET- and BUP-treated individuals would exhibit lower brain MOR availability than healthy controls, (2) BUP would result in greater receptor occupancy than MET due to its higher MOR affinity, and (3) systemic pharmacokinetic measures, indexed by hepatic tracer kinetics, would differentially relate to brain MOR availability across treatments, reflecting distinct degrees of coupling between peripheral and central compartments.

## Methods

### Participants

The study was conducted in accordance with the Declaration of Helsinki, with all procedures approved by the University of Pennsylvania’s Institutional Review Board (Penn IRB) and the study was registered on clinicaltrials.gov (NCT05528848). We recruited 23 English-speaking individuals aged 18-50 (13 HCs and 10 OUD patients, 5 maintained on BUP and 5 on MET) who gave informed consent to participate in the study (Table 1). Prospective participants were first screened by telephone through a partial HIPAA waiver and, if eligible, were seen in person for informed consent and evaluation.

**Table 1.** Demographic and clinical characteristics of healthy controls (HCs) at baseline, methadone-treated OUD patients (MET-OUD) and buprenorphine-treated OUD patients (BUP-OUD). For blocking information, see [9].

| Group | Baseline HCs | MET-OUD | BUP-OUD | P-value* (Overall) | P-value* (MET vs BUP) |
| --- | --- | --- | --- | --- | --- |
| Sample size | 13 | 5 | 5 |  |  |
| Sex (Female/<br>Male) | 6/7 | 0/5 | 1/4 | 0.138 | 0.292 |
| Age (yr) | 29.1 ± 8.3<br>(19.0 - 43.0) | 38.0 ± 2.0<br>(36.0 - 40.0) | 39.6 ± 5.5<br>(33.0 - 47.0) | **0.012** | 0.555 |
| Weight (kg) | 74.5 ± 16.7<br>(56.8 - 120.0) | 105.1 ± 20.5<br>(88.1 - 140.6) | 83.2 ± 11.6<br>(72.2 - 102.1) | **0.009** | **0.071** |
| Body Mass Index | 24.8 ± 4.0<br>(19.4 - 33.6) | 32.0 ± 4.5<br>(27.2 - 38.5) | 27.9 ± 5.4<br>(22.9 - 35.3) | **0.018** | 0.229 |
| Mean MOUD Daily Dose (mg) and MME Dose (mg) | N/A | 158 ± 44.5<br>1185 ± 333.9 | 19.2 ± 3.9<br>1,536 ± 350.5 | N/A | 0.144 |
| Injection Dose (MBq) | 167.1 ± 79.6<br>(73.6 - 313.6) | 158.6 ± 51.6<br>(78.4 - 219.3) | 167.1 ± 79.6<br>(73.6 - 313.6) | 0.617 | 0.314 |

\*Chi-square for count data and ANOVA for continuous measures; MOUD=medication for OUD; MME=mean morphine equivalent dose

| Group | Baseline HCs | MET-ODU | BUP-ODU | P-value*<br>(Overall) | P-value*<br>(MET vs BUP) |
| --- | --- | --- | --- | --- | --- |
| Sample size | 13 | 5 | 5 |  |  |
| Sex (Female/<br>Male) | 6/7 | 0/5 | 1/4 | 0.138 | 0.292 |
| Age (yr) | 29.1 ± 8.3<br>(19.0 - 43.0) | 38.0 ± 2.0<br>(36.0 - 40.0) | 39.6 ± 5.5<br>(33.0 - 47.0) | **0.012** | 0.555 |
| Weight (kg) | 74.5 ± 16.7<br>(56.8 - 120.0) | 105.1 ± 20.5<br>(88.1 - 140.6) | 83.2 ± 11.6<br>(72.2 - 102.1) | **0.009** | **0.071** |
| Body Mass Index | 24.8 ± 4.0<br>(19.4 - 33.6) | 32.0 ± 4.5<br>(27.2 - 38.5) | 27.9 ± 5.4<br>(22.9 - 35.3) | **0.018** | 0.229 |
| Mean MOUD Daily<br>Dose (mg) and<br>MME Dose (mg) | N/A | 158 ± 44.5<br>1185 ± 333.9 | 19.2 ± 3.9<br>1,536 ± 350.5 | N/A | 0.144 |
| Injection Dose<br>(MBq) | 167.1 ± 79.6<br>(73.6 - 313.6) | 158.6 ± 51.6<br>(78.4 - 219.3) | 167.1 ± 79.6<br>(73.6 - 313.6) | 0.617 | 0.314 |

OUD patients met Diagnostic and Statistical Manual of Mental Disorders, 5^th^ edition (DSM-5) criteria [25] for a lifetime diagnosis of OUD and were maintained on a stable dosage of BUP or MET treatment for at least four weeks prior to the screening visit. We excluded individuals with a current severe substance use disorder (SUD) other than OUD or cannabis use disorder, self-reported heavy drug use in the past 3 months, and/or current pharmacological treatment for an SUD other than OUD that could interfere with study procedures or outcomes.

Prospective HCs were excluded if they had a lifetime psychiatric or substance use disorder; were taking psychotropic medications or were pregnant or planning to become pregnant; were breastfeeding; or had a history of epilepsy, brain tumor, brain injury, or material in the body that contraindicates magnetic resonance imaging (MRI).

All participants underwent urine drug testing at both the baseline visit and on the day of PET imaging to ensure that they had not recently used a substance that could interfere with testing procedures. Whereas we reported the total-body effects of naloxone on HCs using [^11^C]CFN and a long axial field-of-view (LAFOV) PET instrument [9], we focus here on MOR availability in OUD patients treated with BUP or MET, with the HCs serving as a comparison group.

### Neuroimaging

#### [^11^C]CFN PET/CT, MRI data acquisition

Participants received a T1-weighted anatomic MRI scan (3-T Prisma Fit, Siemens) prior to the PET scan. Synthesis of [^11^C]CFN was performed using a modified method published by Blecha et al. [26] and is discussed in [9]. All [^11^C]CFN PET scans were conducted on the PennPET Explorer [27, 28]. A low-dose CT scan was performed for anatomical localization and attenuation correction followed by a 90-min dynamic PET. [^11^C]CFN was delivered intravenously as a bolus (Table 1) over <5 sec and was immediately followed by a 30-cc normal saline flush. Venous blood sampling was performed to measure radiometabolites at approximately 10, 15, and 30 mins post injection. As previously discussed [9], an intravenous bolus of naloxone (13 mcg/kg) was delivered over <5 sec 10 to 15 minutes prior to the [^11^C]CFN PET acquisition. PET images were reconstructed using a time-of-flight list-mode ordered subsets expectation maximization (OSEM, 25 subsets) reconstruction algorithm [28]. The reconstructed images had a matrix size of 300×300×712 and a voxel size of 2×2×2 mm^3^.

#### Brain Image analysis

A detailed description of brain analysis methods is in our prior publication [9]. Briefly, a perfusion phase (1-10 min) PET image was cropped to the brain and used for frame-by-frame motion correction followed by registration to the T1 image. T1 and associated PET images were then normalized to Montreal Neurologic Institute (MNI) space [29]. Bilateral VOIs for five, high-MOR expression regions were delineated primarily using the Automated Anatomical Labeling (AAL) atlas [30]: bilateral amygdala, caudate, putamen, thalamus, and ventral tegmentum. The VOI was determined manually for the ventral tegmentum. The distribution volume ratio (DVR) for each brain VOI was computed using the Logan reference tissue model [31] with the visual cortex (modified from calcarine) as the reference tissue. We used a fixed k2’ (0.1237 min^-1^) from the literature [10, 32], as per Dubroff et al. [9]. The DVR was calculated using a 0-90 min time-activity curve (TAC). PET image analyses were performed using PMOD (version 4.2, PMOD Technologies Ltd., Zurich, Switzerland).

Voxel-wise analyses were performed by using Statistical Parametric Mapping 25 (SPM25, Wellcome Centre for Human Neuroimaging, UCL Queen Square Institute of Neurology, London, UK; https://www.fil.ion.ucl.ac.uk/spm)[33] implemented in MATLAB R2025 (MathWorks Inc., Natick, MA). A voxel-wise, two-sample t-test was used to compare the DVR parametric images between HC, MET, and BUP groups. The voxel-wise analyses were evaluated at a significance threshold of 0.0005 with uncorrected statistic and a voxel extent of 50.

We analyzed the findings using a one-way, multivariate analysis of variance (MANOVA). Group (MET-treated, BUP-treated, and baseline HC) was used as the independent factor and DVRs in the 5 regions (ventral tegmentum, thalamus, caudate, putamen, amygdala) were the dependent variables. *A posteriori* Tukey tests were computed between groups. Cohen’s *d* was computed based on the average DVR across the 5 regions for the 3 possible group comparisons (BUP *vs.* baseline HCs, MET *vs.* baseline HCs, BUP *vs.* MET). We report mean and standard deviation of DVR. We used a linear regression to assess the relationship between plasma MET and MOR availability, and a logarithmic model for similar analyses of BUP data.

#### Liver analysis and Vt estimation

VOIs were delineated in the liver and in the descending aorta [9]. Because the liver receives both oxygen-rich blood from the hepatic artery via the celiac trunk (20-25%) and nutrient-rich venous blood from the portal vein derived from the intestines, we used a dual-input, two-tissue compartment model [34] to calculate the volume of distribution (Vt). The plasma input function was derived using a population average parent fraction. At each blood sampling timepoint, group differences in parent fraction of HC, MET, and BUP groups were assessed with a Kruskal Wallis test with an *a posteriori* Dunn’s test. Two one-way MANOVAs were then performed, with group (HCs, HCs with block, MET, BUP) as the independent factor. For each MANOVA [^11^C]CFN liver Vt and whole blood area under the curve (AUC, units standardized uptake value·mins) were the dependent variables. *A posteriori* Tukey tests were computed between groups, and a linear regression was used to assess the relationship between drug plasma level (BUP, MET) and liver Vt.

## Results

### Participants

Table 1 shows the demographics and CFN injection doses for participants with OUD treated with MET (N=5) or BUP (N=5) and HCs scanned both at baseline and with naloxone pretreatment (N=13). Across the three groups, there were significant differences on age (p=0.012), weight (p=0.009), and BMI (p=0.018). *A posteriori* testing showed no significant differences between the two OUD groups, both of which were older and had higher body weight and BMI than HCs. There was a non-significant trend for higher body weight in MET-treated patients than those receiving BUP (p=0.071).

### Brain MOR availability across study groups

In our prior work [9], we found a significant effect on MOR availability of naloxone blockade relative to baseline in HCs (F_1,11_=1132.9, p<0.001). Here, when comparing BUP and MET-treated groups to baseline HCs, we observed differences in MOR availability across groups in all five key brain regions (F_10,34_=5.6, p<0.001, *η*^2^=0.62) (**Figure 1**). Compared to baseline HCs, MOR availability was lower in the BUP-treated patients across all regions (all p<0.001), and in the MET-treated patients in the ventral tegmentum and amygdala (p<0.05). MOR availability was lower in BUP- than MET-treated patients in the thalamus, caudate, putamen, and amygdala (all p<0.05). The average DVR across the 5 regions was lower by 39.9±15.9% (Cohen’s *d*=3.35) in BUP (1.79±0.45) than baseline HCs (2.98±0.22). Similarly, the average DVR was lower in MET (2.55±0.23) than baseline HCs by 14.2±9.9% (Cohen’s *d*=1.90). The BUP DVR was lower by 29.9±18.8% than the MET DVR (Cohen’s *d*=2.14). (See Supplemental Data Table 1 for the individual DVRs.) Parametric DVR maps are shown in **Figure 2**. Beyond the five *a priori* VOIs, voxelwise SPM analysis revealed greater MOR availability in the right middle cingulum, right middle frontal gyrus, and left superior medial frontal gyrus in MET than BUP (**Figure 3**, Supplemental Data Table 2).

**Figure 1.**
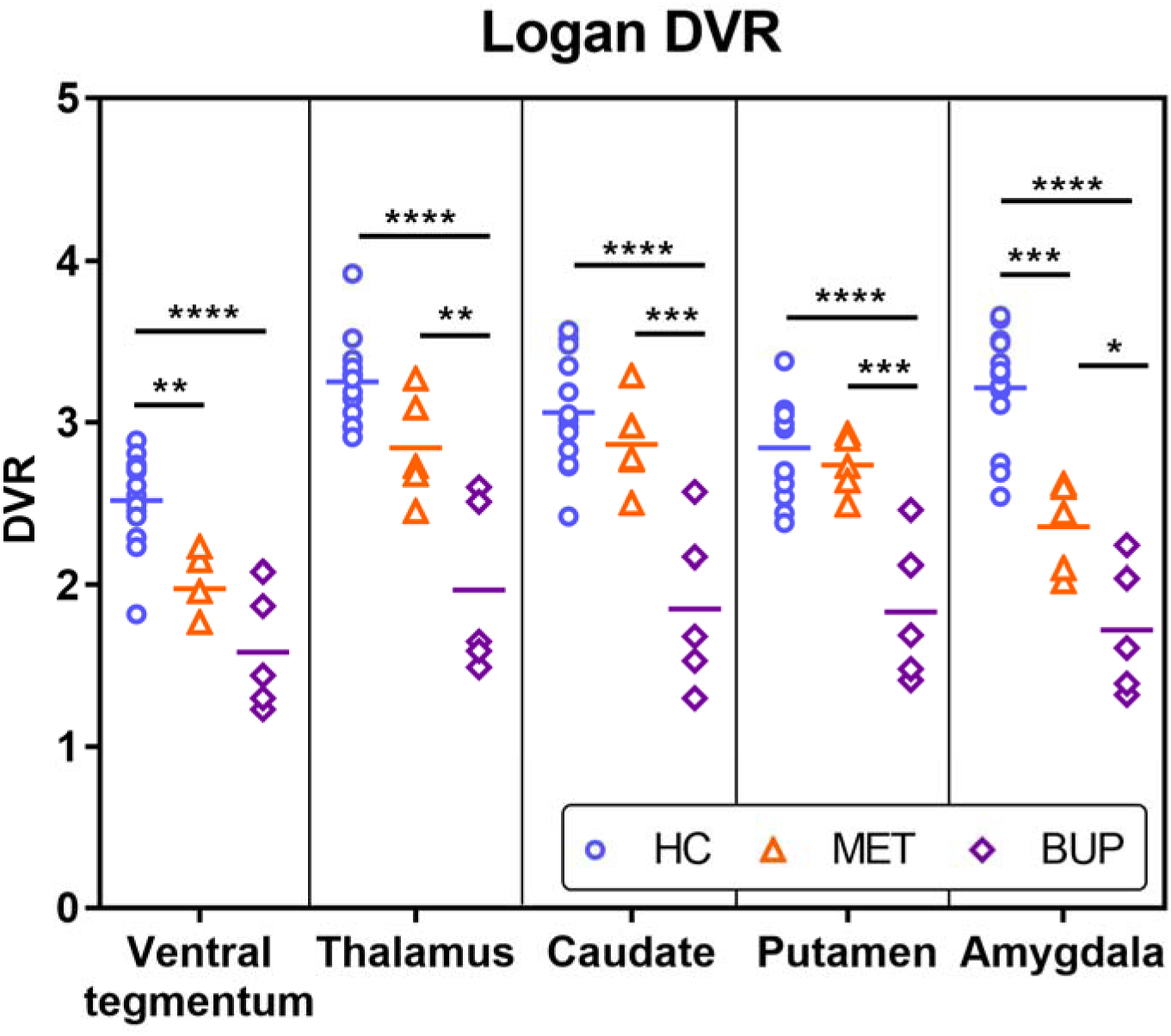
Regional Logan [^11^C]CFN-PET DVR values for each group (HCs, MET, and BUP) across five *a priori* MOR-rich brain regions: ventral tegmentum, thalamus, caudate, putamen, and amygdala.

**Figure 2.**
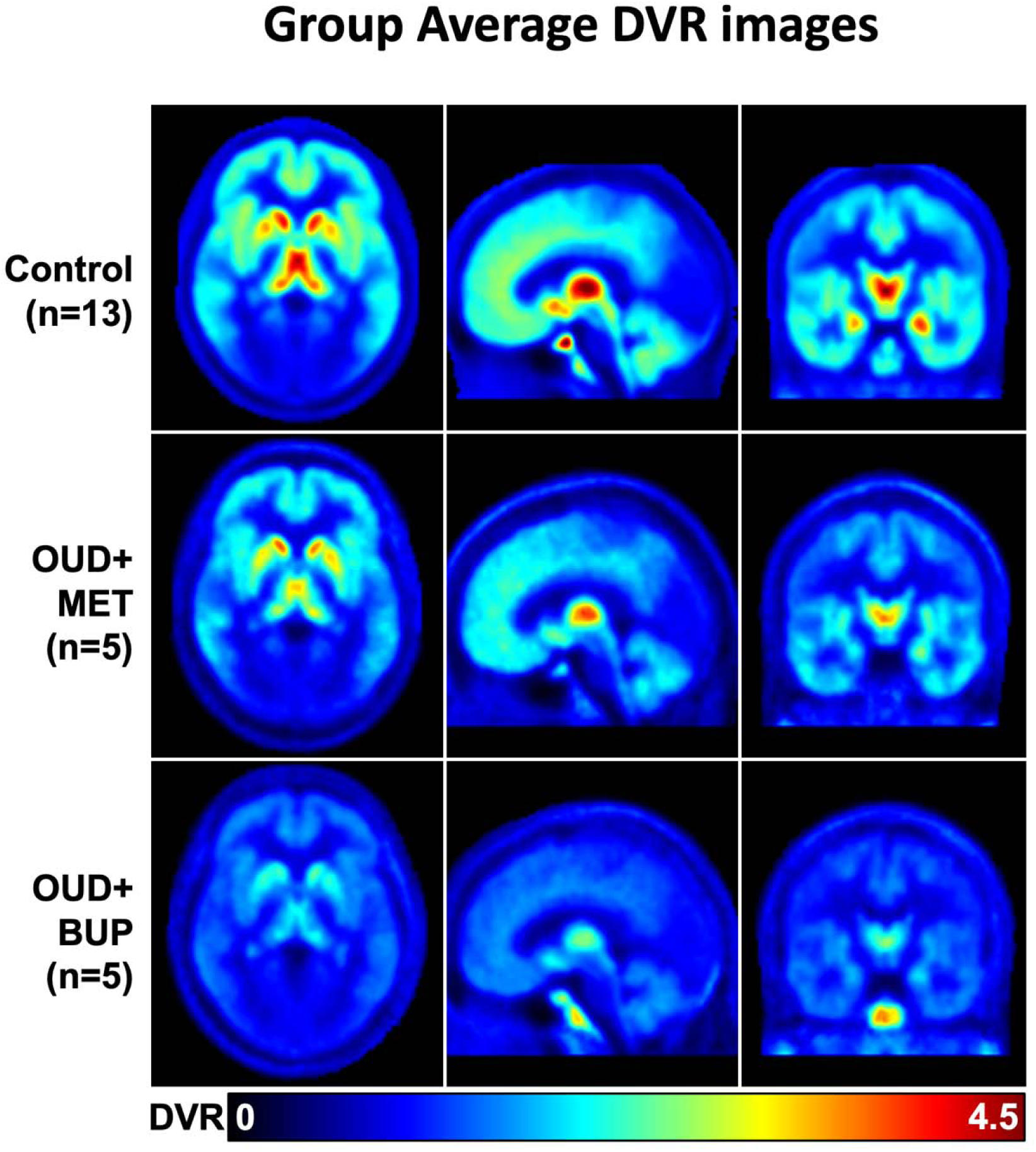
Parametric [^11^C]CFN-PET brain DVR maps in HCs (top row), MET-treated individuals with OUD (middle row), and BUP-treated individuals with OUD (bottom row). Images are displayed in the transaxial (left column), sagittal (middle column), and coronal (right column) planes.

**Figure 3.**
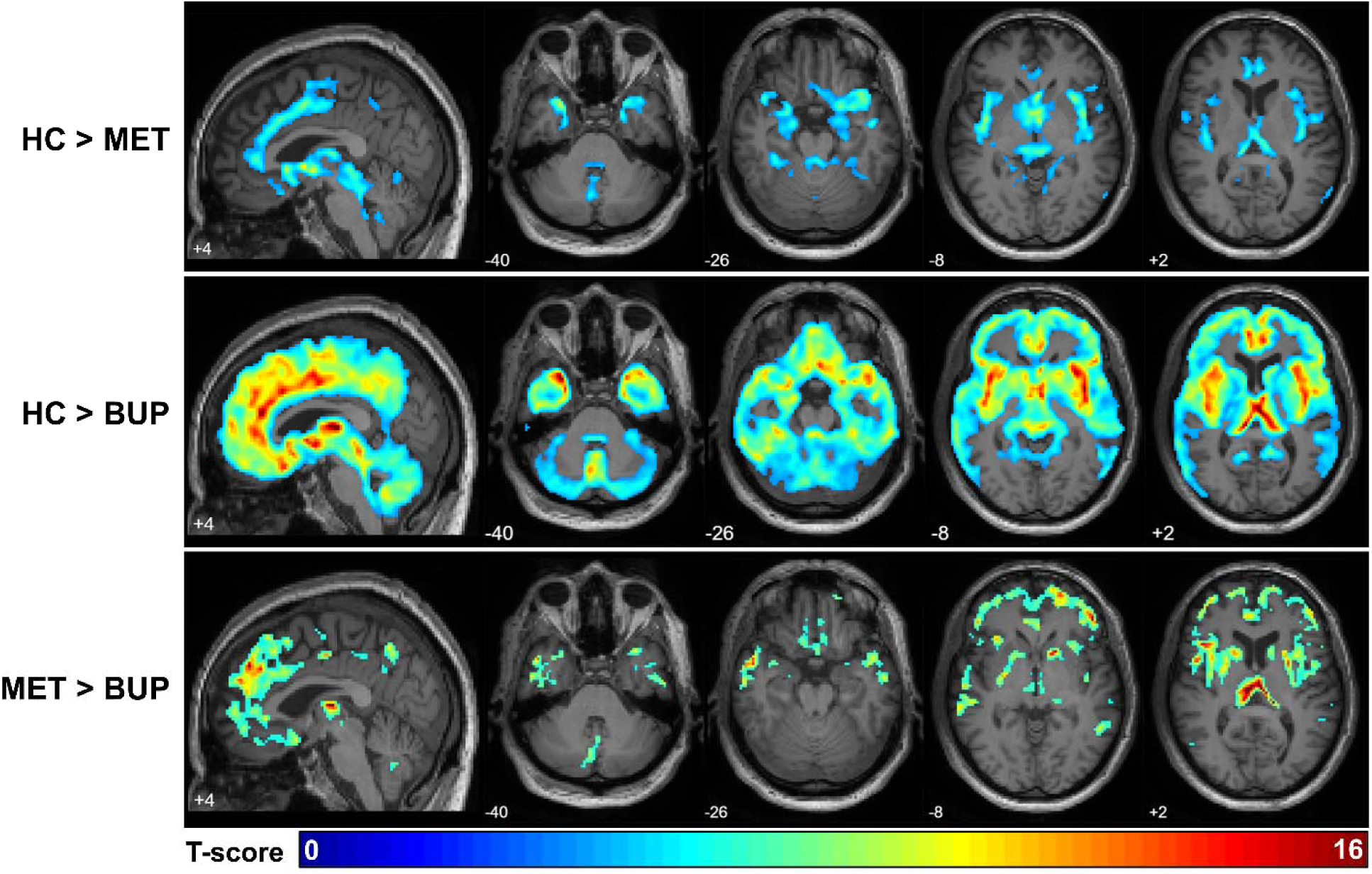
Statistical parametric maps showing pairwise group comparisons of MOR availability. Top row: HCs> MET; Middle row: HCs> BUP; Bottom row: MET> BUP. Images are shown in the sagittal plane (column 1) and transaxial plane (column 2-4). BUP is associated with widespread reductions in MOR availability, whereas MET shows more regionally circumscribed effects. No signal is observed in occipital regions due to negligible MOR expression.

### Whole blood AUC radiometabolite differences across study groups

Individual parent fractions and implemented population parent fraction are included in supplemental data. There were no statistical differences in parent fraction at 10- and 30-min post-injection. However, at 20 min, parent fractions differed among groups (p=0.03), with Dunn’s test showing a difference between MET and BUP (p<0.01). There was a significant difference in whole blood AUC by single-factor MANOVA (p<0.001). The AUC of HCs was similar at baseline (53.7 ± 6.8) and after naloxone pretreatment (49.5 ± 6.5). Higher, more variable values of AUC were seen in both the MET (67.5 ± 24.9) and BUP (73.3± 9.2) groups. The difference between HCs at baseline and with naloxone blockade was not significant via Tukey’s test. However, there was a significant difference in AUC when comparing as vs BUP (p<0.01), HCs with naloxone vs BUP (p<0.01), and HCs with naloxone vs MET (p<0.03).

### Liver Vt and association with brain MOR availability across study groups

Hepatic Vt differed across experimental groups by a one-way MANOVA (p<0.03), which compared baseline HCs (24.3±5.1), HCs with naloxone (23.6±4.5), MET (17.4±4.6), and BUP (16.6 ±2.3), as shown in **Figure 4**. Tukey’s test revealed no difference between HCs baseline and HCs with naloxone, but significant differences when comparing baseline HCs vs MET (p<0.04), baseline HCs vs BUP (p<0.02), and HCs with naloxone vs BUP (p<0.03). There was a strong linear correlation between liver Vt and brain MOR availability in MET (R^2^=0.68), though the correlation was weak in BUP (R^2^=0.30). No correlation was observed in HCs at baseline. These findings are shown in **Figure 5**.

**Figure 4.**
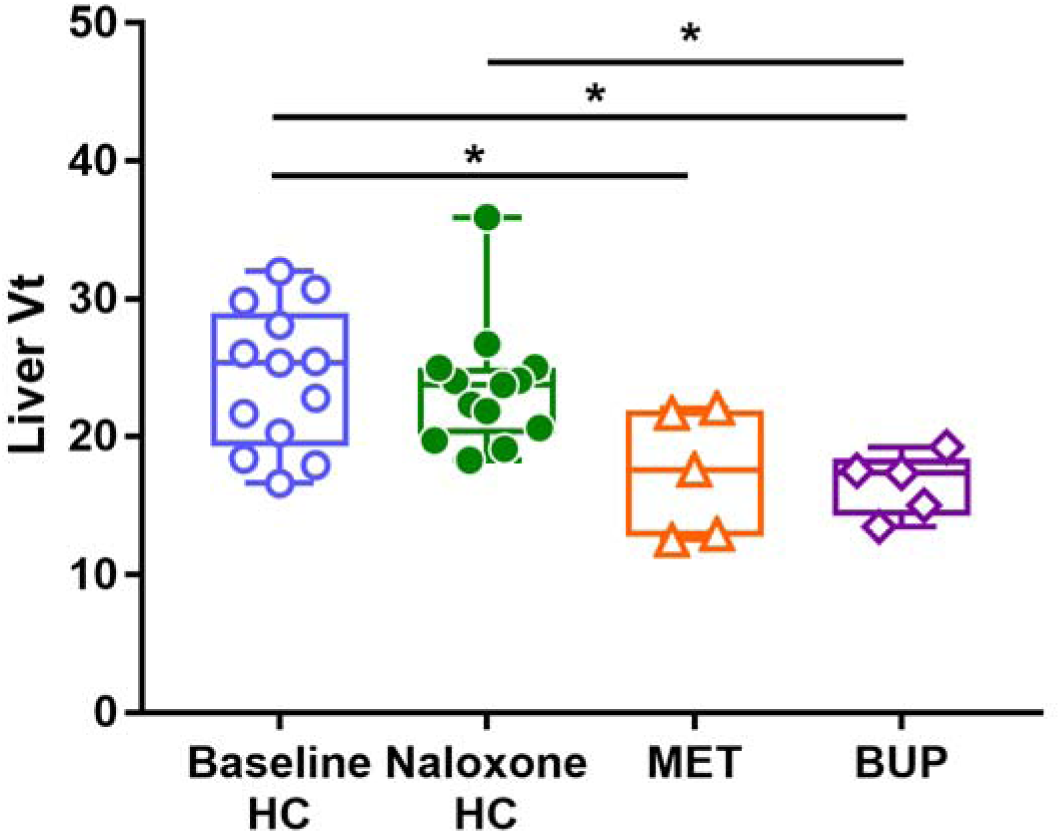
[^11^C]CFN Liver Vt across 4 experimental groups: HCs baseline vs HCs with naloxone block vs MET vs BUP (single factor MANOVA p<0.03). Tukey’s test revealed no difference between HCs baseline and HCs with naloxone. However, significant differences were observed when comparing baseline HCs vs MET (p<0.04), baseline HCs vs BUP (p<0.02), and HCs with naloxone vs BUP (p<0.03).

**Figure 5.**
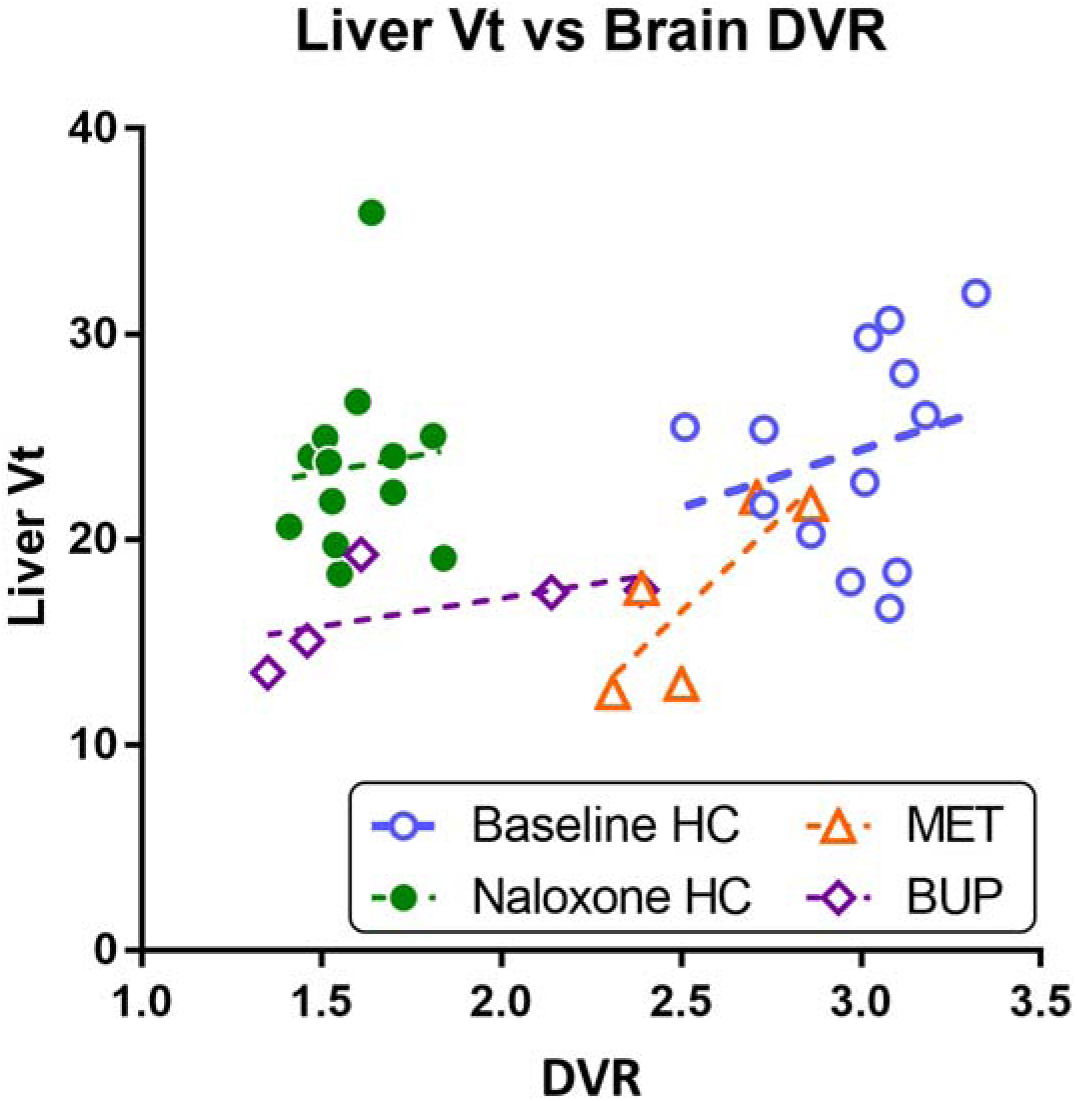
[^11^C]CFN Liver Vt vs [^11^C]CFN DVR mean of 5 MOR rich regions: ventral tegmentum, thalamus, caudate, putamen, and amygdala shows a strong correlation in MET (R^2^=0.68), a weak correlation in BUP (R^2^=0.30), and no correlation in HCs.

### Associations between MET and BUP serum levels in patients with OUD

**Figure 6** shows an inverse linear relationship between MET serum level and mean brain MOR availability (Y=-0.0008219*X+3.305, R^2^=0.83, p=0.03) and an inverse logarithmic relationship between BUP level and MOR availability (R^2^=0.96). Liver Vt showed a strong inverse correlation with MET serum levels (R^2^=0.63) but none with BUP serum levels.

**Figure 6.**
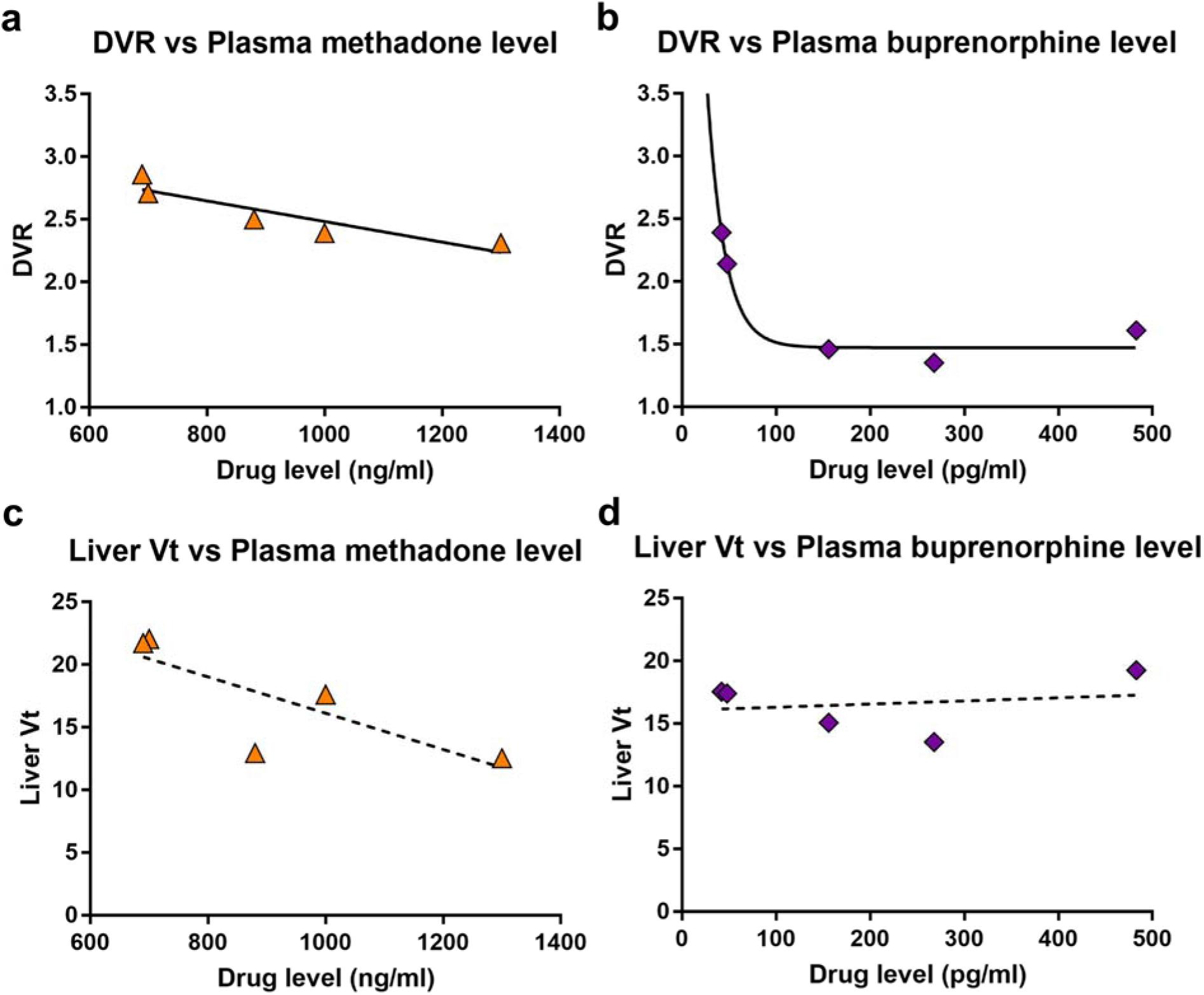
Mean [^11^C]CFN DVR of the five brain regions (ventral tegmentum, thalamus, caudate, putamen, and amygdala) illustrates a linear inverse relationship between MET level and DVR (a) and an inverse logarithmic relationship between BUP level and DVR (b). [^11^C]CFN liver Vt showed an inverse linear relationship with MET serum drug level (c) but not in BUP (d)

## Discussion

Using high-sensitivity [¹¹C]CFN PET imaging on a total-body system, we found that individuals with OUD treated with either MET or BUP exhibited substantially lower MOR availability than HCs. However, MOR availability was significantly lower in BUP- than MET-treated patients in four of the five brain regions examined. All observed effects were very large in magnitude (all Cohen’s *d*>1.90). These differences align with the distinct pharmacodynamic profiles of the medications: BUP’s high MOR affinity and partial agonist effects result in more sustained receptor engagement, whereas MET—a full agonist with lower MOR affinity—may engage fewer receptors at steady-state serum levels [12, 14].

The inverse relationship between serum drug levels and MOR availability further supports these pharmacologic differences. The linear association among MET-treated patients reflects a direct dose-dependent response. The logarithmic relationship among BUP-treated patients indicates that even low serum levels produce a high level of occupancy, consistent with BUP’s high affinity and known ceiling effect [35]. These findings may explain why relatively modest doses of BUP effectively suppress withdrawal, despite having a lower intrinsic activity than MET.

Although MET is a MOR full agonist, receptor availability was significantly reduced only in the ventral tegmentum and amygdala, regions central to reward processing and affect regulation. In contrast, BUP-treated individuals showed widespread reductions in MOR availability, including in the thalamus, caudate, and putamen. This broader engagement may reflect BUP’s sustained occupancy across circuits implicated in reward, stress, and habit formation. These differences in regional receptor availability may contribute to clinical differences seen with the two medications. Taken together, these findings suggest preserved coupling between systemic exposure, hepatic tracer kinetics, and central MOR availability for MET, whereas BUP shows no relationship with receptor-level occupancy at administered drug levels. MET’s selective action in key affective regions may contribute to greater capacity than BUP to retain individuals in opioid agonist treatment, as observed in meta-analyses [5, 36].

Our findings partially replicate those of Kling et al. [15], who reported 19–32% lower opioid receptor availability in the thalamus, amygdala, caudate, anterior cingulate cortex, and putamen of long-term MET-treated individuals using [¹^8^F]cyclofoxy PET. Similarly, we observed significantly lower MOR availability in the amygdala and caudate, among other regions, of MET-treated patients. However, our use of [¹¹C]CFN, a selective MOR agonist tracer, rather than a mixed mu/kappa ligand, yielded a more specific assessment of MOR availability. Here, we extend the findings of Kling et al. [15] by demonstrating that BUP is associated with broader and more substantial reductions in MOR availability than MET, findings that may reflect differences in receptor affinity and occupancy profiles. Although we focused on five *a priori* VOIs with the greatest MOR availability to detect differences across experimental groups, SPM analysis revealed greater MOR availability in the right middle cingulum and right middle frontal and left superior medial frontal gyri in MET-treated than BUP-treated OUD patients (**Figure 3**, Supplemental Data Table 2). Whereas dysfunction in the cingulate and prefrontal cortices has been implicated in addictive disorders, manifesting as impaired executive function and greater opioid cue reactivity [37], the greater treatment retention seen with MET than BUP could reflect a differential impact on these circuits among patients receiving these treatments.

While there was no difference among HCs between the baseline and naloxone scans, aortic AUCs differed significantly for BUP relative to baseline in HCs, and for both BUP and MET relative to HCs with naloxone pretreatment. BUP-treated patients also had a lower hepatic Vt and a higher AUC than baseline HCs. Importantly, hepatic Vt should be interpreted as a pharmacokinetic marker of systemic tracer distribution and clearance rather than hepatic receptor density, providing an index of total-body [¹¹C]CFN availability. Within this framework, hepatic Vt reflects systemic ligand availability that contributes to the fraction of tracer available for central receptor binding.

A key finding of this study is that hepatic tracer kinetics were systematically related to brain MOR availability in a treatment-dependent manner. In MET-treated patients, hepatic Vt showed a strong inverse relationship with serum MET levels and a robust association with brain MOR availability, consistent with a relatively linear systemic-central pharmacokinetic-pharmacodynamic coupling. In contrast, BUP-treated patients, showed a decoupling of MOR occupancy and peripheral pharmacokinetics. Finally, BUP-treated patients showed lower brain DVR, indicating greater overall receptor occupancy than in MET-treated patients. However, larger studies that directly assess (1) liver pharmacokinetics of MET and BUP with [¹¹C]CFN, (2) measurement of [¹¹C]CFN plasma radiometabolites in OUD patients, and (3) a wider range of therapeutic MET and BUP dose levels are needed to refine our understanding of these systemic-central coupling relationships.

The use of a long axial field-of-view and high sensitivity PET scanner in this study enabled us to detect differences in MOR availability in small, subcortical regions such as the ventral tegmentum, which are often inaccessible in standard PET systems, while simultaneously imaging the liver. This new generation of PET technology enhances spatial resolution, sensitivity, and axial field of view, offering novel opportunities for mapping neuroreceptor systems in addiction treatment research both in the central nervous system and the periphery [9, 27].

Based on these findings, [¹¹C]CFN PET imaging may be useful in individualizing OUD treatment. For example, identifying the serum level at which maximal MOR occupancy is present using [¹¹C]CFN PET could inform dose titration, which could then be readily translated to clinical care. Moreover, distinct regional MOR occupancy patterns may eventually serve as neurobiological markers to guide opioid agonist selection, particularly for patients with co-occurring conditions (e.g., mood disorders, obesity) that may interact with opioid receptor systems, though such an approach would require wider availability of PET imaging capabilities than currently exists.

Limitations of this study include its cross-sectional design and small sample size, though the latter was mitigated by the very large size of the effects seen. Additionally, interindividual variability in dosing, drug metabolism, receptor density/expression, comorbid conditions, radiotracer parent fraction, and the presence of radiometabolites in the liver could influence the quantification of MOR availability and liver Vt. Thus, evaluation of the impact of these subject-specific factors on MOR availability and liver Vt measured using [¹¹C]CFN PET is needed to apply these finding to individual therapeutic strategies. We also found statistical differences between plasma fractions at 20 mins post-injection. While the differences in these data may be attributed to the presence of outliers and noisy measurements, additional studies may benefit from radiometabolite correction at the treatment group-specific or individual level. Furthermore, the OUD patient sample was predominantly male, which limits the potential to generalize the findings to females treated with MET or BUP. Thus, larger, longitudinal studies in which samples are balanced on sex are needed to determine whether early or sustained MOR occupancy predicts such key outcomes as treatment retention, opioid abstinence, or affective stability.

Future work should also compare the association of MOR availability with clinical traits such as hedonic tone, anhedonia, drug craving, and weight gain across different opioid agonist treatments. Incorporating multimodal imaging, e.g., combining PET with fMRI or DTI, could inform receptor-level changes that translate to alterations in network-level function. Ultimately, this line of research may support more personalized and mechanistically informed approaches to OUD treatment.

## Conclusions

To the best of our knowledge this study is the first study to 1) directly compare MOR availability using [^11^C]CFN total-body PET between BUP- and MET-treated individuals with OUD and 2) associate MOR receptor availability with systemic pharmacokinetic measures. We demonstrate that both opioid medications are associated with lower MOR availability than HCs, with more widespread receptor engagement observed in BUP-treated patients. We also show a drug-specific, liver-brain pharmacokinetic profile. MET exhibits a linear relationship between systemic exposure, hepatic tracer distribution, and central receptor availability, whereas BUP shows decoupling. In this work, we used total-body [^11^C]CFN PET imaging to investigate the potential link between peripheral drug disposition and central receptor engagement *in vivo*. Since a drug-specific relationship between systemic pharmacokinetics and central MOR function was observed, additional total-body PET studies may be useful for improving the mechanistic understanding of opioid agonist treatments in OUD.

## Data Availability

All data produced in the present work are contained in the manuscript.

## Author Contribution

Corinde Wiers, Robert Mach, Hsiagoju Lee, Henry Kranzler, and Jacob Dubroff contributed to the study conception and design. All authors contributed to carrying out of the studies. Elizabeth Li, Chia-Ju Hsieh, Corinde Wiers, and Jacob Dubroff contributed to data analysis. The first draft of the manuscript was written by Henry Kranzler and Jacob Dubroff. All authors commented on previous versions of the manuscript. All authors read and approved the final manuscript.

## Acknowledgements

The authors thank the University of Pennsylvania Physics and Instrumentation Group for assistance in conducting the PennPET Explorer imaging studies.

## Declarations

### Ethics approval

The study was conducted in accordance with the Declaration of Helsinki, with all procedures approved by the University of Pennsylvania’s Institutional Review Board (Penn IRB #851051) and the study was registered on clinicaltrials.gov (NCT05528848).

Informed consent Informed consent was obtained from all individual participants included in this study. Prospective participants were first screened by telephone through a partial HIPAA waiver and, if eligible, were seen in person for informed consent and evaluation.

## References

1. CDC. Understanding the Opioid Overdose Epidemic. U.S. Centers for Disease Control and Preventoin; 2025.

2. Robert M, Jouanjus E, Khouri C, Fouilhe Sam-Lai N, Revol B. The opioid epidemic: A worldwide exploratory study using the WHO pharmacovigilance database. Addiction. 2023;118:771–5. doi:10.1111/add.16081.

3. Volkow ND, Blanco C. Fentanyl and Other Opioid Use Disorders: Treatment and Research Needs. Am J Psychiatry. 2023;180:410–7. doi:10.1176/appi.ajp.20230273.

4. Blanco C, Volkow ND. Management of opioid use disorder in the USA: present status and future directions. Lancet. 2019;393:1760–72. doi:10.1016/S0140-6736(18)33078-2.

5. Mattick RP, Breen C, Kimber J, Davoli M. Buprenorphine maintenance versus placebo or methadone maintenance for opioid dependence. Cochrane Database Syst Rev. 2014;2014:CD002207. doi:10.1002/14651858.CD002207.pub4.

6. Meyer M, Strazdins E, Guessoum A, Westenberg JN, Appenzeller-Herzog C, Cattaneo M, et al. Relative risks of adverse effects across different opioid agonist treatments-A systematic review and meta-analysis. Addiction. 2025;120:1112–26. doi:10.1111/add.70000.

7. Enns BG-A, B.C.; Min, J.E.; Carter, A.; Siebert, U.; Nosyk, B. Population-Level Health Benefits and Harms Associated With Buprenorphine/Naloxone vs Methadone. JAMA Netw Open. 2025;8:e2551337. doi:10.1001/jamanetworkopen.2025.51337.

8. Nosyk B, Min JE, Homayra F, Kurz M, Guerra-Alejos BC, Yan R, et al. Buprenorphine/Naloxone vs Methadone for the Treatment of Opioid Use Disorder. JAMA. 2024;332:1822–31. doi:10.1001/jama.2024.16954.

9. Dubroff JG, Hsieh CJ, Wiers CE, Lee H, Schmitz A, Li EJ, et al. [(11)C]Carfentanil PET Whole-Body Imaging of mu-Opioid Receptors: A First in-Human Study. J Nucl Med. 2025. doi:10.2967/jnumed.124.269413.

10. Frost JJ, Douglass KH, Mayberg HS, Dannals RF, Links JM, Wilson AA, et al. Multicompartmental analysis of [11C]-carfentanil binding to opiate receptors in humans measured by positron emission tomography. J Cereb Blood Flow Metab. 1989;9:398–409. doi:10.1038/jcbfm.1989.59.

11. Greenwald M, Johanson CE, Bueller J, Chang Y, Moody DE, Kilbourn M, et al. Buprenorphine duration of action: mu-opioid receptor availability and pharmacokinetic and behavioral indices. Biol Psychiatry. 2007;61:101–10. doi:10.1016/j.biopsych.2006.04.043.

12. Greenwald MK, Johanson CE, Moody DE, Woods JH, Kilbourn MR, Koeppe RA, et al. Effects of buprenorphine maintenance dose on mu-opioid receptor availability, plasma concentrations, and antagonist blockade in heroin-dependent volunteers. Neuropsychopharmacology. 2003;28:2000–9. doi:10.1038/sj.npp.1300251.

13. Hsieh CJ, Hou C, Lee H, Tomita C, Schmitz A, Plakas K, et al. Total-body imaging of mu-opioid receptors with [(11)C]carfentanil in non-human primates. Eur J Nucl Med Mol Imaging. 2024. doi:10.1007/s00259-024-06746-2.

14. Zubieta J, Greenwald MK, Lombardi U, Woods JH, Kilbourn MR, Jewett DM, et al. Buprenorphine-induced changes in mu-opioid receptor availability in male heroin-dependent volunteers: a preliminary study. Neuropsychopharmacology. 2000;23:326–34. doi:10.1016/S0893-133X(00)00110-X.

15. Kling MA, Carson RE, Borg L, Zametkin A, Matochik JA, Schluger J, et al. Opioid receptor imaging with positron emission tomography and [(18)F]cyclofoxy in long-term, methadone-treated former heroin addicts. J Pharmacol Exp Ther. 2000;295:1070–6.

16. Melichar JK, Hume SP, Williams TM, Daglish MR, Taylor LG, Ahmad R, et al. Using [11C]diprenorphine to image opioid receptor occupancy by methadone in opioid addiction: clinical and preclinical studies. J Pharmacol Exp Ther. 2005;312:309–15. doi:10.1124/jpet.104.072686.

17. Erbs E, Faget L, Scherrer G, Matifas A, Filliol D, Vonesch JL, et al. A mu-delta opioid receptor brain atlas reveals neuronal co-occurrence in subcortical networks. Brain Struct Funct. 2015;220:677–702. doi:10.1007/s00429-014-0717-9.

18. Charbogne P, Kieffer BL, Befort K. 15 years of genetic approaches in vivo for addiction research: Opioid receptor and peptide gene knockout in mouse models of drug abuse. Neuropharmacology. 2014;76 Pt B:204-17. doi:10.1016/j.neuropharm.2013.08.028.

19. Hsu DT, Sanford BJ, Meyers KK, Love TM, Hazlett KE, Walker SJ, et al. It still hurts: altered endogenous opioid activity in the brain during social rejection and acceptance in major depressive disorder. Mol Psychiatry. 2015;20:193–200. doi:10.1038/mp.2014.185.

20. Ahmad T, Valentovic MA, Rankin GO. Effects of cytochrome P450 single nucleotide polymorphisms on methadone metabolism and pharmacodynamics. Biochem Pharmacol. 2018;153:196–204. doi:10.1016/j.bcp.2018.02.020.

21. Ferrari A, Coccia CP, Bertolini A, Sternieri E. Methadone--metabolism, pharmacokinetics and interactions. Pharmacol Res. 2004;50:551-9. doi:10.1016/j.phrs.2004.05.002.

22. Kong L, Walz AJ. Metabolism of the active carfentanil metabolite, 4-Piperidinecarboxylic acid, 1-(2-hydroxy-2-phenylethyl)-4-[(1-oxopropyl)phenylamino]-, methyl ester in vitro. Toxicol Lett. 2022;367:32–9. doi:10.1016/j.toxlet.2022.07.006.

23. Oda Y, Mizutani K, Hase I, Nakamoto T, Hamaoka N, Asada A. Fentanyl inhibits metabolism of midazolam: competitive inhibition of CYP3A4 in vitro. Br J Anaesth. 1999;82:900–3. doi:10.1093/bja/82.6.900.

24. Dubroff JG, Hsieh CJ, Wiers CE, Lee H, Schmitz A, Li EJ, et al. [(11)C]Carfentanil PET Whole-Body Imaging of mu-Opioid Receptors: A First in-Human Study. J Nucl Med. 2025;66:1112–8. doi:10.2967/jnumed.124.269413.

25. American Psychiatric Association., American Psychiatric Association. DSM-5 Task Force. Diagnostic and statistical manual of mental disorders : DSM-5. Fifth edition. ed.

26. Blecha JE, Henderson BD, Hockley BG, VanBrocklin HF, Zubieta JK, DaSilva AF, et al. An updated synthesis of [(11) C]carfentanil for positron emission tomography (PET) imaging of the mu-opioid receptor. J Labelled Comp Radiopharm. 2017;60:375–80. doi:10.1002/jlcr.3513.

27. Karp JS, Viswanath V, Geagan MJ, Muehllehner G, Pantel AR, Parma MJ, et al. PennPET Explorer: Design and Preliminary Performance of a Whole-Body Imager. J Nucl Med. 2020;61:136–43. doi:10.2967/jnumed.119.229997.

28. Pantel AR, Viswanath V, Daube-Witherspoon ME, Dubroff JG, Muehllehner G, Parma MJ, et al. PennPET Explorer: Human Imaging on a Whole-Body Imager. J Nucl Med. 2020;61:144–51. doi:10.2967/jnumed.119.231845.

29. Collins DL, Zijdenbos AP, Kollokian V, Sled JG, Kabani NJ, Holmes CJ, et al. Design and construction of a realistic digital brain phantom. IEEE Trans Med Imaging. 1998;17:463–8. doi:10.1109/42.712135.

30. Tzourio-Mazoyer N, Landeau B, Papathanassiou D, Crivello F, Etard O, Delcroix N, et al. Automated anatomical labeling of activations in SPM using a macroscopic anatomical parcellation of the MNI MRI single-subject brain. Neuroimage. 2002;15:273–89. doi:10.1006/nimg.2001.0978.

31. Logan J, Fowler JS, Volkow ND, Wang GJ, Ding YS, Alexoff DL. Distribution volume ratios without blood sampling from graphical analysis of PET data. J Cereb Blood Flow Metab. 1996;16:834–40. doi:10.1097/00004647-199609000-00008.

32. Hirvonen J, Aalto S, Hagelberg N, Maksimow A, Ingman K, Oikonen V, et al. Measurement of central µ-opioid receptor binding in vivo with PET and [^11^C] carfentanil: a test–retest study in healthy subjects. European journal of nuclear medicine and molecular imaging. 2009;36:275–86.

33. Tierney TM, Alexander NA, Avila NL, Balbastre Y, Barnes G, Bezsudnova Y, et al. SPM 25: open source neuroimaging analysis software. arXiv preprint arXiv:250112081. 2025.

34. Zuo Y, Sarkar S, Corwin MT, Olson K, Badawi RD, Wang G. Structural and practical identifiability of dual-input kinetic modeling in dynamic PET of liver inflammation. Phys Med Biol. 2019;64:175023. doi:10.1088/1361-6560/ab1f29.

35. Lutfy K, Cowan A. Buprenorphine: a unique drug with complex pharmacology. Curr Neuropharmacol. 2004;2:395–402. doi:10.2174/1570159043359477.

36. Fareed A, Vayalapalli S, Stout S, Casarella J, Drexler K, Bailey SP. Effect of methadone maintenance treatment on heroin craving, a literature review. J Addict Dis. 2011;30:27–38. doi:10.1080/10550887.2010.531672.

37. Goldstein RZ, Volkow ND. Dysfunction of the prefrontal cortex in addiction: neuroimaging findings and clinical implications. Nat Rev Neurosci. 2011;12:652–69. doi:10.1038/nrn3119.

