## supplemental data for "Total-body [^11^C]carfentanil PET: liver-brain axis in methadone vs. buprenorphine treatment"

Supplemental Data Table 1: Individual [^11^C]CFN DVR estimates

| Group | Participant | DVR Ventral Tegmentum | DVR Thalamus | DVR Caudate | DVR Putamen | DVR Amygdala |
| --- | --- | --- | --- | --- | --- | --- |
| HC | 851051-01 | 2.89 | 3.39 | 3.02 | 2.96 | 3.64 |
| HC | 851051-04 | 2.81 | 3.30 | 2.73 | 2.54 | 3.66 |
| HC | 851051-05 | 2.55 | 3.15 | 2.74 | 2.44 | 2.75 |
| HC | 851051-06 | 2.45 | 2.97 | 3.52 | 3.08 | 3.51 |
| HC | 851051-07 | 2.74 | 3.52 | 2.97 | 2.99 | 3.20 |
| HC | 851051-08 | 2.61 | 3.92 | 3.48 | 3.38 | 3.23 |
| HC | 851051-09 | 2.70 | 3.19 | 3.35 | 3.05 | 3.30 |
| HC | 851051-10 | 2.49 | 3.29 | 3.19 | 3.05 | 3.37 |
| HC | 851051-13 | 2.29 | 3.06 | 2.94 | 2.69 | 2.69 |
| HC | 851051-18 | 1.82 | 3.34 | 3.05 | 2.62 | 3.49 |
| HC | 851051-23 | 2.23 | 2.98 | 2.42 | 2.38 | 2.54 |
| HC | 851051-24 | 2.42 | 2.91 | 3.57 | 3.08 | 3.11 |
| HC | 851051-26 | 2.72 | 3.27 | 2.83 | 2.70 | 3.32 |
| MET | 851051-201 | 2.15 | 3.09 | 2.98 | 2.73 | 2.62 |
| MET | 851051-202 | 2.23 | 3.27 | 3.29 | 2.93 | 2.60 |
| MET | 851051-204 | 1.96 | 2.45 | 2.77 | 2.90 | 2.44 |
| MET | 851051-206 | 1.77 | 2.73 | 2.78 | 2.64 | 2.02 |
| MET | 851051-224 | 1.77 | 2.68 | 2.50 | 2.49 | 2.10 |
| BUP | 851051-200 | 1.23 | 1.49 | 1.30 | 1.41 | 1.32 |
| BUP | 851051-209 | 1.44 | 1.65 | 1.68 | 1.69 | 1.61 |
| BUP | 851051-211 | 1.30 | 1.59 | 1.53 | 1.48 | 1.39 |
| BUP | 851051-217 | 2.08 | 2.60 | 2.57 | 2.46 | 2.24 |
| BUP | 851051-218 | 1.87 | 2.51 | 2.17 | 2.12 | 2.04 |

Supplemental Data Table 2: Statistical Parametric Mapping (SPM) of MET > BUP MOR availability corresponding to Figure 3 (bottom row). 17 regions were identified using a significance level of < 0.0005 with uncorrected statistic and a voxel extent of 50.

| Cluster-level | | | | Peak-level | | | | | Coordinate (mm) | | |  |
| --- | --- | --- | --- | --- | --- | --- | --- | --- | --- | --- | --- | --- |
| P_FWE-corr_ | P_FDR-corr_ | K_E_ | P_uncorr_ | P_FWE-corr_ | P_FDR-corr_ | T | Z_E_ | P_uncorr_ | x | y | z | Region |
| 0.000 | 0.000 | 887.000 | 0.000 | 0.074 | 0.798 | 23.439 | 5.071 | 0.000 | 8 | -18 | 2 | Right Thalamus |
|  |  |  |  | 0.165 | 0.798 | 20.491 | 4.917 | 0.000 | 16 | -28 | -2 | Right Thalamus |
|  |  |  |  | 0.224 | 0.798 | 19.465 | 4.857 | 0.000 | -6 | -22 | 0 | Left Thalamus |
| 0.000 | 0.000 | 15106.000 | 0.000 | 0.304 | 0.798 | 18.488 | 4.796 | 0.000 | -4 | 40 | 40 | Left Superior Medial Frontal |
|  |  |  |  | 0.340 | 0.798 | 18.139 | 4.774 | 0.000 | -2 | 28 | 50 | Left Superior Medial Frontal |
|  |  |  |  | 0.363 | 0.798 | 17.943 | 4.761 | 0.000 | 48 | 40 | -10 | Right Inferior Orbito Frontal cortex |
| 0.000 | 0.000 | 297.000 | 0.000 | 1.000 | 0.798 | 11.144 | 4.165 | 0.000 | 62 | -52 | 14 | Right Middle Temporal |
|  |  |  |  | 1.000 | 0.798 | 10.519 | 4.089 | 0.000 | 66 | -44 | 16 | Right Superior Temporal |
|  |  |  |  | 1.000 | 0.798 | 9.512 | 3.954 | 0.000 | 52 | -58 | 36 | Right Angular |
| 0.001 | 0.000 | 98.000 | 0.000 | 1.000 | 0.798 | 10.956 | 4.143 | 0.000 | -56 | -52 | 16 | Left Middle Temporal |
|  |  |  |  | 1.000 | 0.798 | 9.953 | 4.015 | 0.000 | -58 | -52 | 8 | Left Middle Temporal |
|  |  |  |  | 1.000 | 0.820 | 7.397 | 3.604 | 0.000 | -52 | -62 | 4 | Left Middle Temporal |
| 0.000 | 0.000 | 214.000 | 0.000 | 1.000 | 0.798 | 10.894 | 4.135 | 0.000 | -58 | -34 | 26 | Left SupraMarginal |
|  |  |  |  | 1.000 | 0.798 | 9.356 | 3.931 | 0.000 | -58 | -18 | 22 | Left Postcentral |
|  |  |  |  | 1.000 | 0.798 | 8.751 | 3.840 | 0.000 | -56 | -36 | 34 | Left SupraMarginal |
| 0.019 | 0.002 | 63.000 | 0.000 | 1.000 | 0.798 | 10.714 | 4.113 | 0.000 | 6 | -10 | 52 | Right Middle Cingulum |
|  |  |  |  | 1.000 | 0.799 | 7.971 | 3.710 | 0.000 | 10 | -8 | 60 | Right Supplementary Motor Area |
|  |  |  |  | 1.000 | 0.878 | 6.579 | 3.435 | 0.000 | 8 | -16 | 46 | Right Middle Cingulum |
| 0.013 | 0.002 | 68.000 | 0.000 | 1.000 | 0.798 | 10.520 | 4.089 | 0.000 | -30 | -2 | 58 | Left Precentral |
|  |  |  |  | 1.000 | 0.799 | 7.990 | 3.713 | 0.000 | -30 | -14 | 54 | Left Precentral |
| 0.000 | 0.000 | 351.000 | 0.000 | 1.000 | 0.798 | 10.412 | 4.075 | 0.000 | 0 | -54 | 44 | Left Precuneus |
|  |  |  |  | 1.000 | 0.798 | 9.966 | 4.017 | 0.000 | -2 | -34 | 38 | Left Middle Cingulum |
|  |  |  |  | 1.000 | 0.798 | 9.127 | 3.897 | 0.000 | -4 | -50 | 30 | Left Posterior Cingulum |
| 0.001 | 0.000 | 107.000 | 0.000 | 1.000 | 0.798 | 10.278 | 4.058 | 0.000 | 66 | -24 | 18 | Right Superior Temporal |
|  |  |  |  | 1.000 | 0.798 | 9.791 | 3.993 | 0.000 | 60 | -32 | 14 | Right Superior Temporal |
| 0.016 | 0.002 | 65.000 | 0.000 | 1.000 | 0.798 | 9.836 | 3.999 | 0.000 | 30 | -2 | 58 | Right Middle Frontal |
| 0.003 | 0.000 | 88.000 | 0.000 | 1.000 | 0.798 | 9.765 | 3.989 | 0.000 | 58 | -16 | 28 | Right SupraMarginal |
|  |  |  |  | 1.000 | 0.798 | 8.576 | 3.812 | 0.000 | 52 | -22 | 18 | Right Rolandic operculum |
|  |  |  |  | 1.000 | 0.804 | 7.776 | 3.675 | 0.000 | 46 | -26 | 22 | Right Rolandic operculum |
| 0.001 | 0.000 | 109.000 | 0.000 | 1.000 | 0.798 | 9.755 | 3.988 | 0.000 | -16 | -44 | 50 | Left Precuneus |
|  |  |  |  | 1.000 | 0.798 | 8.605 | 3.816 | 0.000 | -12 | -54 | 60 | Left Precuneus |
|  |  |  |  | 1.000 | 0.798 | 8.392 | 3.782 | 0.000 | -20 | -50 | 60 | Left Superior Parietal |
| 0.003 | 0.001 | 86.000 | 0.000 | 1.000 | 0.798 | 9.696 | 3.980 | 0.000 | 36 | -40 | 50 | Right Inferior Parietal |
|  |  |  |  | 1.000 | 0.799 | 7.988 | 3.713 | 0.000 | 44 | -32 | 52 | Right Postcentral |
| 0.015 | 0.002 | 66.000 | 0.000 | 1.000 | 0.798 | 9.696 | 3.980 | 0.000 | 28 | -52 | -56 | Right Cerebellum 8 |
|  |  |  |  | 1.000 | 0.849 | 6.928 | 3.510 | 0.000 | 18 | -62 | -58 | Right Cerebellum 8 |
|  |  |  |  | 1.000 | 0.886 | 6.532 | 3.425 | 0.000 | 36 | -56 | -54 | Right Cerebellum 8 |
| 0.001 | 0.000 | 102.000 | 0.000 | 1.000 | 0.798 | 9.448 | 3.944 | 0.000 | 54 | -50 | -6 | Right Inferior Temporal |
|  |  |  |  | 1.000 | 0.798 | 8.184 | 3.747 | 0.000 | 60 | -48 | -14 | Right Inferior Temporal |
| 0.000 | 0.000 | 128.000 | 0.000 | 1.000 | 0.798 | 8.686 | 3.829 | 0.000 | 52 | -32 | -2 | Right Middle Temporal |
|  |  |  |  | 1.000 | 0.811 | 7.602 | 3.643 | 0.000 | 60 | -34 | -4 | Right Middle Temporal |
|  |  |  |  | 1.000 | 0.868 | 6.690 | 3.460 | 0.000 | 62 | -26 | -12 | Right Middle Temporal |
| 0.004 | 0.001 | 84.000 | 0.000 | 1.000 | 0.798 | 8.382 | 3.780 | 0.000 | -4 | -68 | -40 | Left Cerebellum 8 |
|  |  |  |  | 1.000 | 0.799 | 8.000 | 3.715 | 0.000 | 2 | -58 | -40 | Vermis 9 |
|  |  |  |  | 1.000 | 0.813 | 7.526 | 3.628 | 0.000 | -8 | -80 | -38 | Left Cerebellum Crus2 |

**Figure S1: Parent fraction across baseline, MET, and BUP**

Parent fraction of [^11^C]CFN across 3 experimental groups: HCs baseline, MET, and BUP. Measurements taken at approximately 10, 20, and 30 min post-injection.

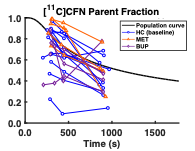
